# From Epidemic to Endemic: Longitudinal Surveillance of Congenital Zika Syndrome in Brazil

**DOI:** 10.64898/2026.07.07.26357442

**Authors:** Raiane Oliveira Ferreira, Hui Ling Ma, Patrícia Pestana Garcez, Mayana Zatz

**Author notes:** Corresponding author: Mayana Zatz, PhD, Human Genome and Stem Cell Research Center (HUG-CELL), Institute of Biosciences, University of São Paulo, 055080-090, Cidade Universitária, São Paulo, Brazil.

## Abstract

Zika virus (ZIKV) emerged in Brazil in 2015, causing an unprecedented epidemic of Congenital Zika Syndrome (CZS). A decade later, longitudinal analyses evaluating temporal trends and subnational heterogeneity in CZS burden remain limited. Using publicly available data from SINAN/DATASUS and the RESP-Microcephaly registry (SVS/Ministry of Health, updated July 2024), we conducted a descriptive ecological analysis of ZIKV infection and CZS in Brazil from 2015 to 2023. Of 331,309 notified Zika cases (2015-2023), 213,350 occurred in 2016, followed by an 91.75% decline in 2017 and sustained low-level endemic circulation thereafter. Among 3,751 confirmed microcephaly cases, 1,828 were confirmed with ZIKV etiology. The Northeast region accounted for 75.4% of confirmed cases despite representing approximately 27% of the national population. State-level analyses revealed distinct epidemiological patterns, including persistent microcephaly notifications of non-Zika etiology in Minas Gerais and continued detection of ZIKV-attributed CZS in Amazonas and Goiás through 2023. These findings highlight pronounced geographic disparities in congenital Zika burden, reflect significant heterogeneity in diagnostic capacity, and underscore the need for sustained surveillance and systematic etiological investigation of congenital abnormalities in the post-epidemic era.

**AUTHOR SUMMARY:** When Zika virus spread across Brazil in 2015, a wave of babies was born with abnormally small heads and severe brain damage, a condition now known as Congenital Zika Syndrome. The epidemic shocked the world, but by 2017 case numbers had dropped sharply, leading many to ask: has this syndrome disappeared? In this study, we analyzed eight years of Brazilian public health data (2015–2023) to track Zika infections and cases of congenital brain abnormalities over time and across all Brazilian states. Our findings reveal that, while the dramatic epidemic peak has passed, Zika virus continues to circulate at low levels and babies continue to be born with Zika-related brain damage each year. We also uncovered striking geographic inequalities: the Northeast region, home to about 27% of Brazil’s population, accounted for over 75% of confirmed cases. States with higher poverty and poorer sanitation were hardest hit. We found that the ability to correctly identify the cause of brain abnormalities varied widely across states, raising concerns that some cases may go undetected. Our results highlight that Congenital Zika Syndrome has not disappeared, it has simply become less visible. Sustained monitoring and investigation of congenital brain abnormalities remain essential public health priorities.

## INTRODUCTION

Since its first isolation in a sentinel rhesus macaque in the Zika Forest of Uganda in 1947, Zika virus (ZIKV) has remained a neglected pathogen for several decades, with only sporadic cases of human infection reported across sub-Saharan Africa and Southeast Asia [1–4]. This profile shifted with the 2007 outbreak on Yap Island, Federated States of Micronesia, the first confirmed epidemic outside Africa, followed by a large outbreak in French Polynesia in 2013–2014, during which the first associations with Guillain-Barré syndrome were reported [5]. The subsequent introduction of ZIKV into the Americas, confirmed in northeastern Brazil in early 2015, triggered a public health crisis of unprecedented scale [6, 7]. Brazil was disproportionately affected, driven by a combination of high Aedes aegypti vector density, rapid and heterogeneous urbanization, marked inequalities in basic sanitation coverage, and a fully susceptible population with no prior immunological exposure to ZIKV. In adults, infection is typically asymptomatic or manifests as a mild, self-limiting febrile illness characterized by rash, conjunctivitis, arthralgia, and low-grade fever. In contrast, the Brazilian outbreak, revealed a previously unrecognized consequence of ZIKV infection: an abrupt and geographically clustered increase in congenital neurological abnormalities temporally associated with maternal infection during pregnancy, first identified by clinicians in 2015 [8].

Subsequent investigations confirmed the neurotropism of ZIKV and its capacity to infect and disrupt neural progenitor cells during critical windows of fetal neurodevelopment [9–11]. Although microcephaly emerged as the most visible manifestation, it represents only a subset of a broader spectrum of congenital injury, defined in 2016 by the World Health Organization as Congenital Zika Syndrome (CZS), which encompasses intracranial calcifications, ventriculomegaly, corpus callosum abnormalities, cerebellar hypoplasia, ocular malformations, and long-term neurodevelopmental impairment, including severe motor and cognitive disability [12]. The recognition of CZS as a distinct clinical entity represented a critical epidemiological milestone, demonstrating that arboviral infections can cause catastrophic teratogenic outcomes [13]. It also underscored that disease burden is not randomly distributed, but disproportionately concentrated in populations with limited access to antenatal care, fragile surveillance systems, and structural poverty, a pattern whose underlying drivers remain incompletely understood.

A decade after the emergence of ZIKV in Brazil, reported incidence has declined substantially from the 2015–2016 epidemic peak; however, transmission persists at low endemic levels, and CZS notifications continue to accrue annually. To date, most published epidemiological analyses have focused on the acute epidemic phase or the immediate clinical sequelae of congenital infection [14, 15]. This leaves a critical gap: ecological analyses conducted at the national or regional level aggregate heterogeneous transmission dynamics and may conceal anomalous patterns in localized anomalies that may hold independent epidemiological significance. Furthermore, many existing studies rely on absolute case counts without standardization for population size, which can distort the geographic distribution of disease burden in a country as demographically heterogeneous as Brazil.

In this study, we conducted a temporal, geographical, and demographic analysis of ZIKV infection and CZS in Brazil from 2015 to 2023, using publicly available national surveillance data. By evaluating incidence patterns across regions and population subgroups, we aimed to characterize the first decade of CZS and identify regional heterogeneity in the distribution of ZIKV-associated congenital outcomes. This approach provides an updated overview of the long-term epidemiological patterns of CZS following the introduction of ZIKV into the Americas, with particular attention to spatial disparities that may be obscured in aggregated analyses.

## METHODOLOGY

### Study design

We conducted a retrospective epidemiological analysis of Zika virus infection and CZS in Brazil from 2015 to 2023, using secondary data from the Brazilian national public health surveillance system. A descriptive, ecological, time-series approach was applied to quantify and characterize temporal trends, spatial heterogeneity, and demographic patterns in ZIKV incidence and associated congenital outcomes at the subnational level.

### Data sources

All data were retrieved from publicly accessible Brazilian Ministry of Health databases. Two primary sources were used:

(1) The Sistema de Informação de Agravos de Notificação (SINAN), accessed via the Tabnet platform (http://tabnet.datasus.gov.br), was the primary source for probable Zika case counts, deaths, and incidence data. SINAN records compulsory notifications of infectious diseases submitted by municipal and state health services throughout Brazil. Data were extracted by year of notification, federative unit of residence, sex, age group, and final classification (probable/confirmed). The extraction was performed for the period from epidemiological week 1 of 2015 through epidemiological week 47 of 2023. (2) The Registro de Eventos em Saúde Pública – Microcefalia (RESP-Microcephaly), also maintained by the Ministry of Health Secretariat of Health Surveillance (SVS), was used for data on CZS notifications. RESP-Microcephaly is a dedicated registry that records suspected and confirmed cases of CZS reported across all Brazilian states. Data were retrieved for the period 2015–2023, stratified by state of residence and year of notification, as available at the date of the last platform update (July 2024). The national aggregate and state-level disaggregated data were both extracted to enable regional analysis.

### Data availability

All data used in this study are publicly available through the DATASUS Tabnet platform (http://tabnet.datasus.gov.br) and the RESP-Microcephaly registry of the Brazilian Ministry of Health Secretariat of Health Surveillance (SVS), updated July 2024.

## RESULTS

### Temporal trends in ZIKV notifications, confirmed cases, and diagnostic confirmation rates

Between 2016 and 2023, 279,503 suspected ZIKV cases were reported to the Brazilian National Notifiable Diseases Information System (SINAN), of which 175,499 (62.8%) were laboratory or clinically confirmed (Figure 1). The national epidemiological profile was overwhelmingly dominated by the 2016 outbreak, when 213,350 cases were notified and 141,766 confirmed, representing 76.3% of all notifications and 80.8% of all confirmed cases recorded throughout the entire study period. This finding highlights the exceptional magnitude and abrupt dissemination of ZIKV during its emergence phase in Brazil.

**Figure 1.**
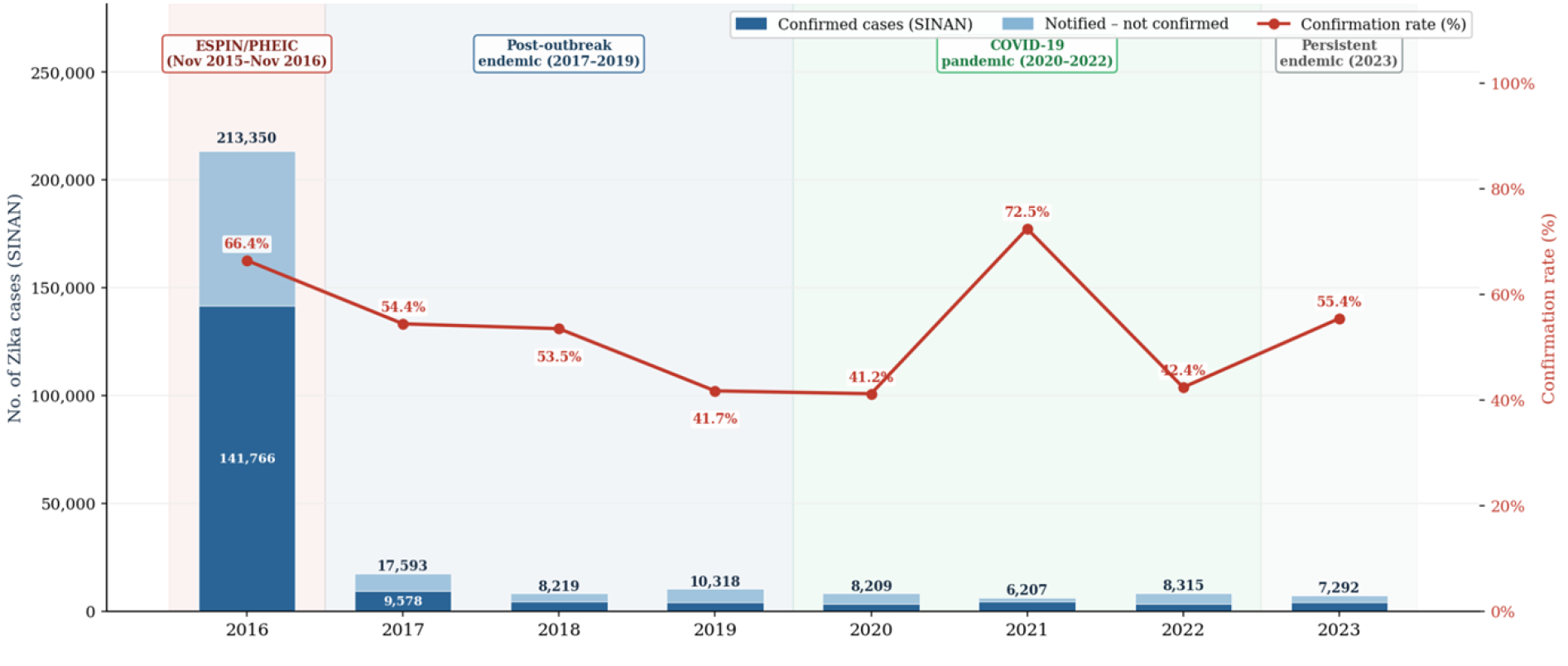
Notified and confirmed ZIKV cases with annual confirmation rate — Brazil, 2016–2023. Stacked bars represent the annual total of notified Zika cases disaggregated into confirmed (dark blue; laboratory or clinical-epidemiological confirmation) and notified-not-confirmed (light blue; notifications without a confirmed final classification). The red line (right axis) represents the annual confirmation rate, calculated as the proportion of notified cases receiving a confirmed classification. Shaded background regions denote four epidemiological periods: ESPIN/PHEIC (national public health emergency and WHO Public Health Emergency of International Concern, November 2015–November 2016); post-outbreak endemic (2017–2019); COVID-19 pandemic (2020–2022); and persistent endemic circulation (2023). Sources: probable cases — SINAN/DATASUS, Epidemiological Bulletin SE 47/2023; confirmed cases — SINAN Tabnet (classification = confirmed), updated 16 April 2024.

Following the epidemic peak, notifications declined sharply to 17,593 cases in 2017 and stabilized thereafter at low endemic levels, ranging from 6,207 cases in 2021 to 10,318 in 2019. Despite this substantial reduction, ZIKV transmission persisted throughout the study period, indicating sustained endemic circulation rather than interruption. The post-epidemic decline likely reflects the accumulation of population immunity after widespread viral exposure, although concurrent changes in surveillance sensitivity, healthcare-seeking behavior, and vector dynamics may also have contributed.

Annual confirmation rates exhibited substantial variability, ranging from 41.2% in 2020 to 72.5% in 2021. Notably, lower confirmation rates were consistently observed during 2019 (41.7%), 2020 (41.2%), and 2022 (42.4%), including periods overlapping with the COVID-19 pandemic. Confirmation rates did not track with notification volume, suggesting that diagnostic ascertainment was influenced not only by transmission intensity but also by operational surveillance factors such as laboratory availability, testing prioritization, and case investigation capacity.

The highest confirmation rate occurred in 2021, coinciding with the lowest number of notified cases. This inverse pattern may reflect more selective notification practices and intensified diagnostic scrutiny during the pandemic period, alongside expanded molecular testing capacity driven by the national COVID-19 response. Conversely, periods of healthcare system strain and prioritization of SARS-CoV-2 surveillance may have reduced the sensitivity of arboviral case notification.

Overall, these findings delineate the transition of ZIKV in Brazil from a large-scale epidemic to persistent endemic transmission, characterized by substantial temporal variability in diagnostic confirmation and surveillance performance.

### Confirmed microcephaly cases and proportion attributed to ZIKV etiology (2015–2023)

A total of 3,762 microcephaly cases were classified as confirmed in the RESP-Microcephaly registry during the study period, of which 1,828 (48.6%) were attributed to ZIKV infection (Figure 2). The ESPIN/PHEIC period (2015–2016) concentrated the overwhelming majority of the burden, with 926 confirmed cases in 2015 and 1,932 in 2016, together representing 76% of all confirmed microcephaly cases over the eight-year series.

**Figure 2:**
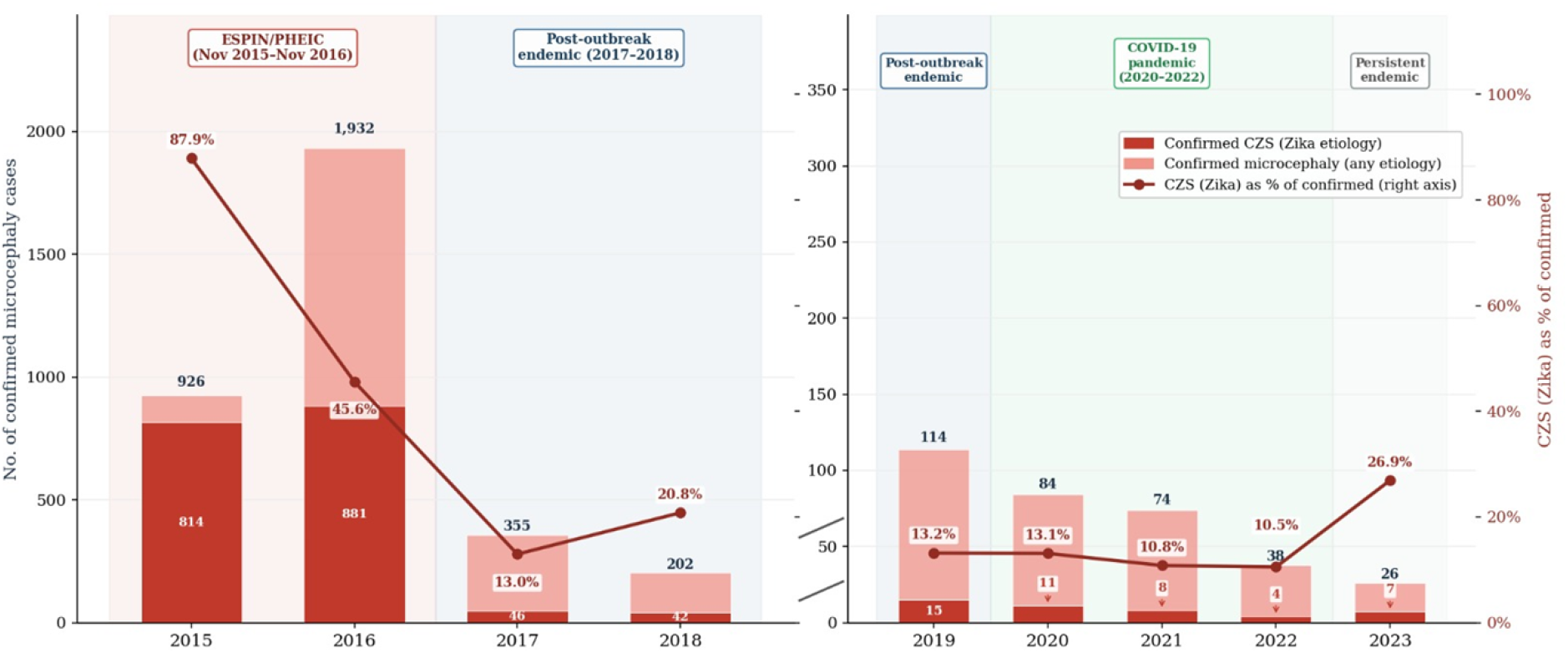
Confirmed microcephaly cases by etiology and proportion attributed to ZIKV — Brazil (2015–2023). Stacked bars represent annual confirmed microcephaly cases disaggregated into confirmed Congenital Zika Syndrome (CZS; ZIKV etiology, dark red) and confirmed microcephaly of any etiology (light red). Values above bars indicate annual totals of confirmed microcephaly; values inside or adjacent to the dark segment indicate confirmed CZS case counts. The dark red line (right axis) represents the annual proportion of confirmed microcephaly cases attributed to ZIKV etiology. The left panel (2015–2018) and right panel (2019–2023) use independent y-axis scales to improve visibility of the smaller case counts recorded in the endemic period. Shaded background regions denote four epidemiological periods: ESPIN/PHEIC (national public health emergency and WHO Public Health Emergency of International Concern, November 2015–November 2016); post-outbreak endemic (2017–2019); COVID-19 pandemic (2020–2022); and persistent endemic circulation (2023). Sources: RESP-Microcephaly / Secretariat of Health Surveillance (SVS) / Ministry of Health, Brazil. Confirmed microcephaly: classification = confirmed, all etiologies (n = 3,762). Confirmed CZS: classification = confirmed AND etiology = ZIKV (n = 1,828). Updated July 2024.

The proportion of microcephaly attributable to ZIKV varied substantially over time, revealing a clear epidemiological transition. In 2015, 814 of 926 confirmed cases (87.9%) were classified as CZS, indicating that ZIKV was the dominant driver of the sudden increase in congenital neurological abnormalities during the initial outbreak phase, particularly in high-transmission settings such as the Northeast region [16]. In 2016, although the absolute number of confirmed CZS cases peaked (n=881), the proportional contribution of ZIKV declined to 45.6%, with more than half of cases attributed to other or undetermined causes. This shift likely reflects the progressive expansion and refinement of surveillance protocols, including the transition from a narrow microcephaly-based definition to a broader spectrum of congenital central nervous system abnormalities. As surveillance intensified, detection of non-ZIKV etiologies likely increased, while the geographic spread of the epidemic into lower-transmission regions may have further diluted the relative contribution of ZIKV.

Following the epidemic peak, the number of confirmed microcephaly cases declined progressively, from 355 in 2017 to 26 in 2023, a 92.7% reduction over six years. This sustained decrease is consistent with reduced population susceptibility after widespread viral circulation, leading to lower maternal exposure and fewer congenital infections. Despite this decline, the proportion of cases attributed to ZIKV remained relatively stable between 2017 and 2022, fluctuating between 10% and 21%, suggesting that most cases during the endemic phase reflect the underlying baseline burden of congenital abnormalities rather than epidemic transmission dynamics.

The pattern observed in 2023 requires cautious interpretation. Of 26 confirmed microcephaly cases, 7 (26.9%) were attributed to ZIKV, the highest proportional contribution since 2018. However, given the small absolute numbers, this increase is unlikely to indicate renewed epidemic transmission. Instead, it may reflect selective case ascertainment and more complete etiological investigation in a context of low notification volume. Nevertheless, together with the continued low-level circulation of ZIKV observed in SINAN surveillance data, these findings reinforce the importance of sustained etiological investigation of congenital abnormalities in the post-epidemic period.

### Subnational distribution of CZS: from notification burden to confirmed Zika etiology

State-level analyses revealed substantial geographic heterogeneity in both the burden of notified microcephaly and the attribution of cases to ZIKV infection. When all confirmed microcephaly cases were considered regardless of etiology (Figure 3), the highest cumulative burdens were observed in Bahia (587 cases) and Pernambuco (471), followed by Rio de Janeiro (318), Minas Gerais (228), Paraíba (227), and São Paulo (199). Overall, the Northeast region concentrated the majority of cases (2,212 notifications), reinforcing its role as the principal epicenter of the congenital manifestations during the Brazilian ZIKV epidemic.

**Figure 3.**
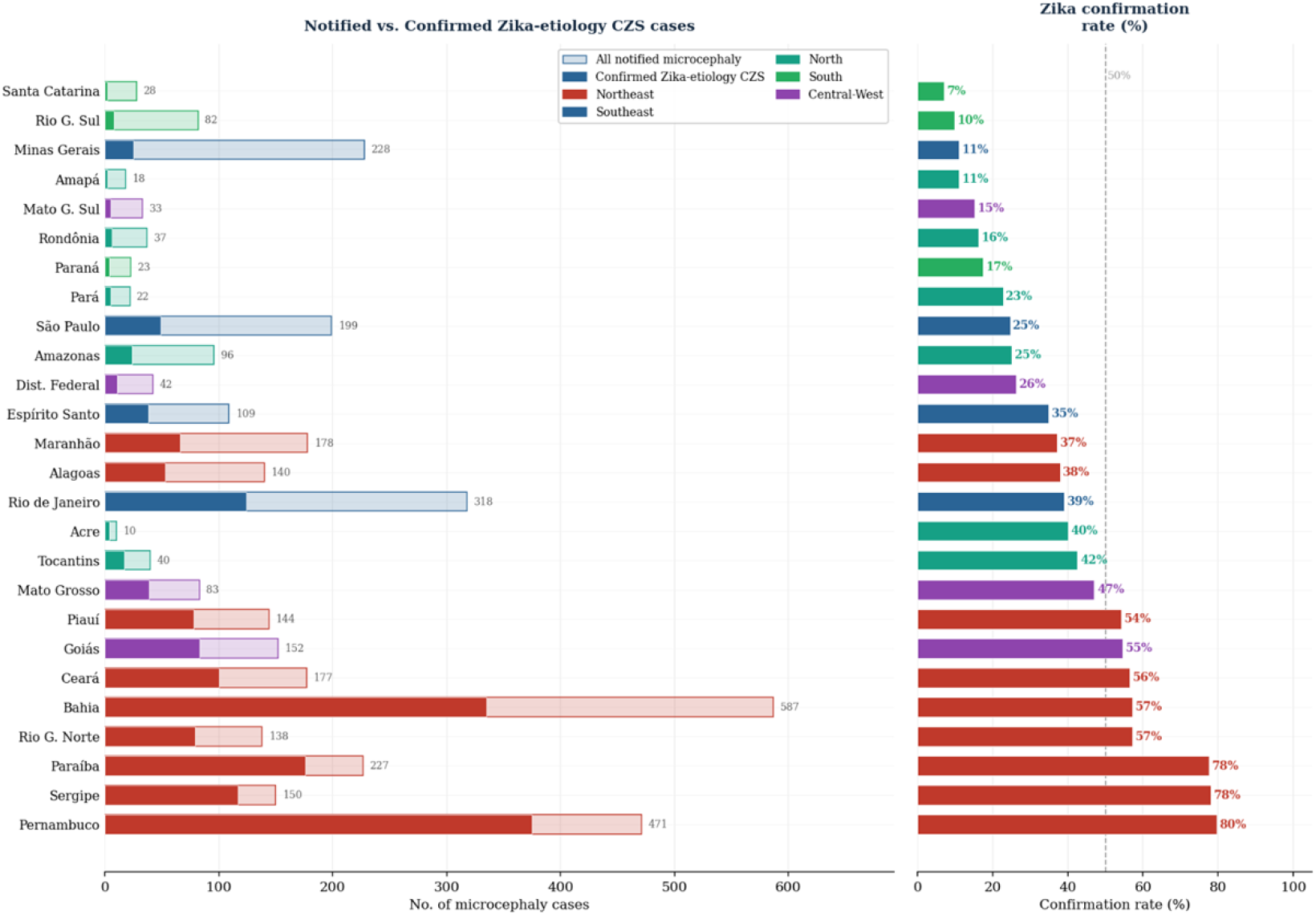
Proportion of notified microcephaly cases confirmed as CZS by state — Brazil, 2015–2023 (cumulative). Left panel: absolute case counts per state; bars represent all notified microcephaly cases regardless of classification or etiology (transparent fill) and confirmed CZS with ZIKV as the attributed etiology (solid fill; n=1,828 nationally). Values inside bars indicate confirmed CZS counts; values to the right indicate total notified cases. Right panel: Zika confirmation rate, calculated as the proportion of notified microcephaly cases confirmed with ZIKV etiology per state. The dashed vertical line denotes the 50% threshold. States are ordered by a descending confirmation rate. Bar colors indicate geographic region. Source: RESP-Microcephaly / Secretariat of Health Surveillance (SVS) / Ministry of Health, Brazil. Updated July 2024.

Restricting the analysis to cases with confirmed ZIKV etiology (Figure 4; n = 1,828) revealed a distinct spatial redistribution. Pernambuco emerged as the leading state in cumulative confirmed CZS burden (375 cases), surpassing Bahia (335), while Paraíba (176) and Rio de Janeiro (124) remained among the most affected. The Northeast accounted for 75.4% of all confirmed CZS cases (1,379/1,828), highlighting the persistent geographic concentration of ZIKV-associated congenital burden even after diagnostic refinement.

**Figure 4.**
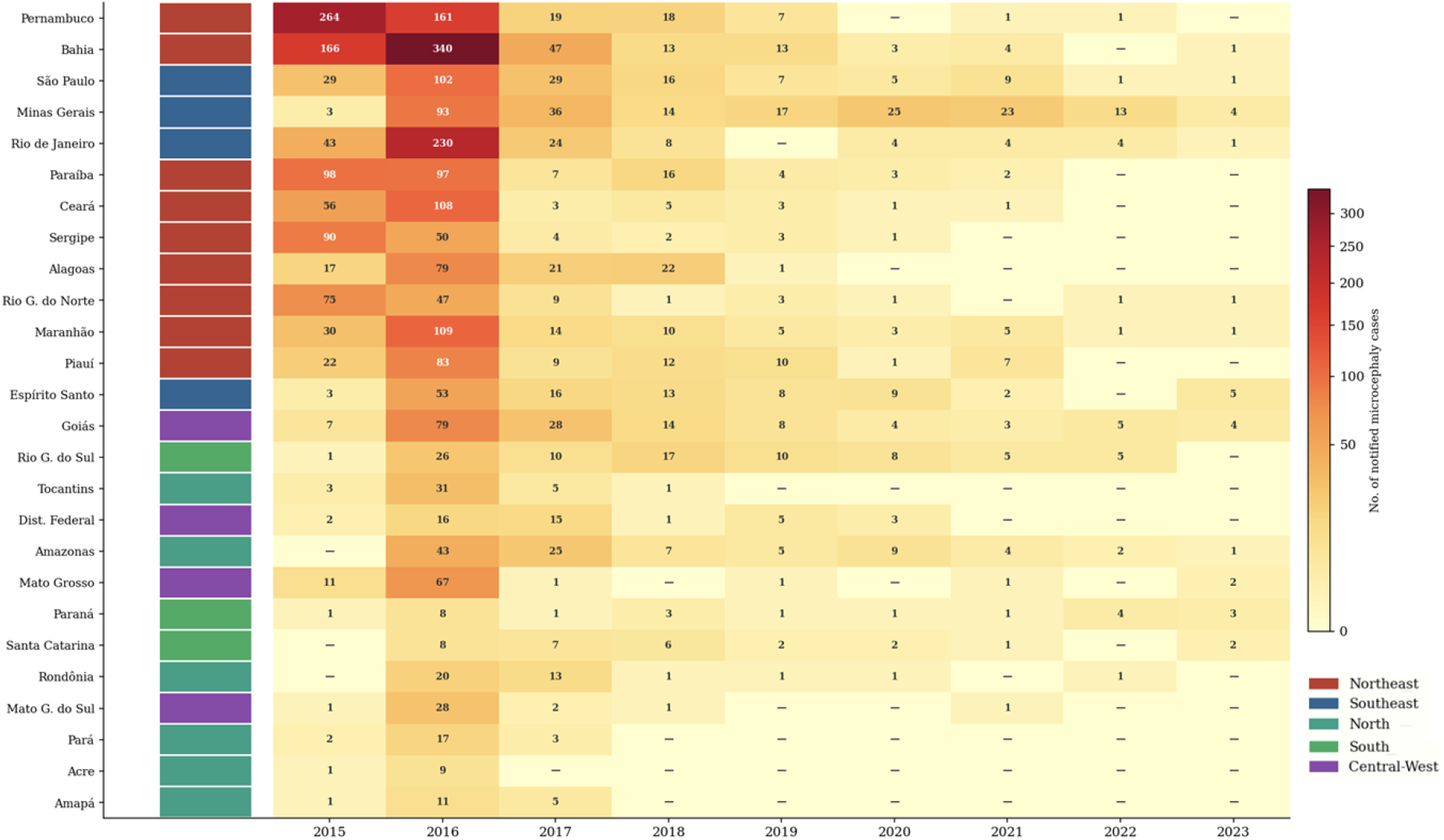
Confirmed microcephaly cases by state — heatmap, Brazil, 2015–2023. Each cell represents the annual count of confirmed microcephaly cases for a given state, regardless of final classification or attributed etiology. Colour intensity is proportional to case count using a power-normalized scale (γ = 0.45) to improve visual discrimination at low case counts. The vertical colour strip on the left indicates the geographic region of each state (red = Northeast; blue = Southeast; teal = North; green = South; purple = Central-West). Values of zero are displayed as “–”. Source: RESP-Microcephaly / SVS / Ministry of Health, Brazil. All notified microcephaly cases regardless of classification or etiology. Updated July 2024.

Comparison of notification burden and confirmed ZIKV attribution demonstrated marked interstate variation in etiological confirmation, synthesized in Figure 3 as cumulative state-level confirmation rates. Nationally, the proportion of notified microcephaly cases attributed to ZIKV ranged from 79.6% in Pernambuco to 7.1% in Santa Catarina. With the exception of Goiás, all states exceeding a 50% confirmation rate were located in the Northeast, reinforcing the spatial alignment between high transmission intensity during the epidemic and subsequent attribution of congenital outcomes to ZIKV.

Within the Northeast, confirmation rates were not homogeneous. Pernambuco, Sergipe, and Paraíba exhibited high proportions of ZIKV-attributed cases (>75%), suggesting that a substantial share of notified congenital abnormalities during the epidemic period was directly linked to ZIKV infection. In contrast, Maranhão (37.1%) and Alagoas (37.9%) showed lower confirmation rates despite high notification burdens, consistent with greater relative contribution of non-Zika congenital etiologies or differences in diagnostic capacity and case investigation.

Outside the Northeast, confirmation rates declined sharply. Minas Gerais presented one of the largest discrepancies between notified and confirmed ZIKV-attributed cases (228 vs 25; 11.0% confirmation rate), while São Paulo displayed a similar pattern (199 vs 49; 24.6%). These gaps suggest that, in several southeastern states, a considerable proportion of microcephaly cases reflects baseline congenital abnormalities unrelated to ZIKV transmission or cases lacking definitive etiological classification.

Temporal patterns within states further highlight divergence between notification burden and confirmed ZIKV attribution. In Minas Gerais, microcephaly notifications persisted throughout the post-epidemic period (Figure 4), whereas confirmed ZIKV-attributed cases declined to near zero after 2020 (Figure 5), indicating a progressive decoupling of congenital outcomes from ZIKV transmission dynamics. In contrast, Amazonas reported sporadic confirmed CZS cases through 2023, standing out as the only northern state with ZIKV-attributed congenital cases in the final year of the series. Although absolute numbers were small, this persistence may reflect ongoing low-level viral circulation in the Amazon region beyond the main epidemic phase.

**Figure 5.**
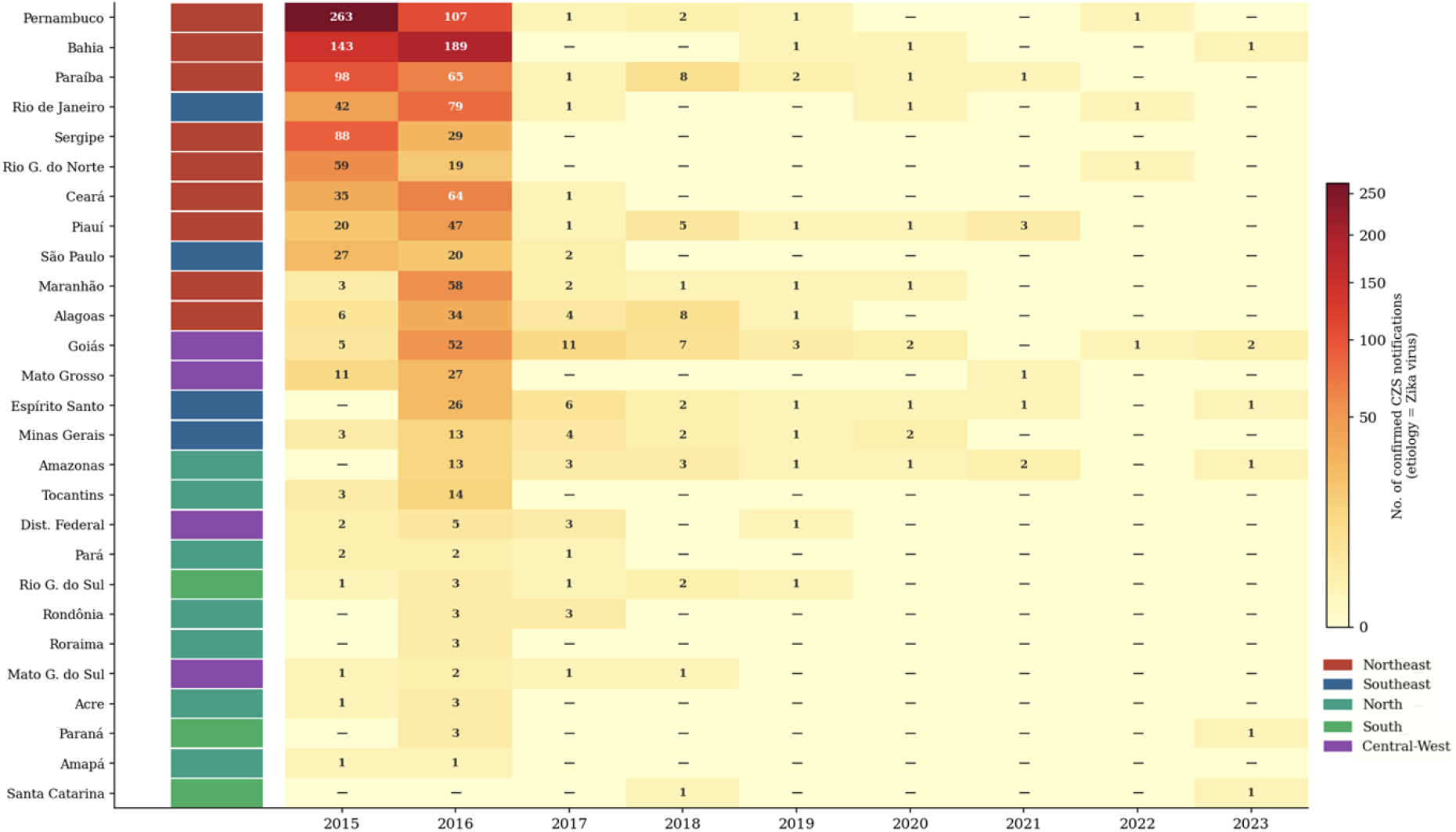
Confirmed Congenital Zika Syndrome (CZS) notifications by state — heatmap, Brazil, 2015–2023. Each cell represents the annual count of confirmed CZS cases attributed to ZIKV (classification = confirmed; etiology = ZIKV) for a given state. Colour intensity is proportional to case count using a power-normalized scale (γ = 0.45). The vertical colour strip on the left indicates the geographic region of each state (red = Northeast; blue = Southeast; teal = North; green = South; purple = Central-West). Values of zero are displayed as “–”. Total confirmed CZS cases with Zika virus etiology: n = 1,828. Source: RESP-Microcephaly / SVS / Ministry of Health, Brazil. Updated July 2024.

In the Central-West region, Goiás exhibited a distinct profile. In addition to presenting the highest cumulative burden of both notified and confirmed cases (152 and 83, respectively), it was the only non-Northeastern state with a confirmation rate exceeding 50% (54.6%) and continued to report confirmed cases through 2023. This pattern suggests sustained vulnerability to ZIKV-associated congenital outcomes outside the historically most affected region.

By contrast, the South region consistently demonstrated the lowest confirmation rates nationwide, Paraná (17.4%), Rio Grande do Sul (9.8%), and Santa Catarina (7.1%), showing minimal attribution of notified microcephaly to ZIKV infection. This pattern aligns with the historically lower intensity of ZIKV transmission in southern Brazil across both epidemic and post-epidemic periods.

## DISCUSSION

A decade after its emergence in Brazil, ZIKV continues to circulate at endemic levels. The findings presented here demonstrate that congenital outcomes associated with prenatal infection have not disappeared; rather, they have become less visible. Although the sustained decline in confirmed CZS notifications following the epidemic peak likely reflects a genuine reduction in transmission intensity, it coincides with a progressive attenuation of diagnostic vigilance outside emergency settings. The persistence of confirmed ZIKV-attributed CZS cases in Amazonas and Goiás through 2023, together with the continued burden of microcephaly notifications in states such as Minas Gerais, largely driven by non-ZIKV or undetermined etiologies, suggests that congenital neurological abnormalities remain an active epidemiological signal requiring systematic investigation, rather than a residual imprint of a resolved epidemic.

The pronounced interstate variation in ZIKV confirmation rates, ranging from 79.6% in Pernambuco to 7.1% in Santa Catarina, cannot be attributed solely to differences in viral transmission. These disparities are closely intertwined with the structural inequalities that shaped the epidemic trajectory. Pernambuco, which presented the highest confirmation rate, is characterized by higher levels of poverty, lower sanitation coverage, and greater Aedes aegypti density, whereas Santa Catarina, at the opposite end of the spectrum, has one of the highest Human Development Indexes in the country and historically limited arboviral transmission. Consistent with this pattern, previous spatial analyses have demonstrated significant associations between the burden of CZS and socioeconomic and health indicators, including spatial correlation and clustering of vulnerability [16]. Together, these findings suggest that the burden of CZS arises from the interaction between biological transmission dynamics and entrenched social vulnerability.

Importantly, the states most affected by ZIKV were often those with the greatest constraints in healthcare access, laboratory infrastructure, and longitudinal follow-up capacity. As new birth cohorts accumulate without prior immunological exposure to ZIKV, the risk of congenital injury in the context of renewed transmission should be viewed not as a historical concern, but as a continuing prospective threat. Sustained ZIKV surveillance, coupled with systematic etiological investigation of all cases of microcephaly and congenital central nervous system abnormalities, therefore remains a critical public health priority for the coming decade.

## Data Availability

No data was generated by this study. The following existing data sources were used: The Sistema de Informação de Agravos de Notificação (SINAN), accessed via the Tabnet platform (http://tabnet.datasus.gov.br) and The Registro de Eventos em Saúde Pública – Microcefalia (RESP-Microcephaly), accessed via resp.saude.gov.br/microcefalia.

http://tabnet.datasus.gov.br

## ACKNOWLEDGMENTS

This study was supported by the São Paulo Research Foundation (FAPESP, grants 2013/08028-1 and 2014/50931-3) and by INCT (Instituto Nacional de Ciência e Tecnologia, grant 465355/2014). The authors also acknowledge the fellowships from FAPESP granted to Raiane Oliveira Ferreira (2020/14109-8) and Hui Ling Ma (2022/02212-4).

## REFERENCES

[1] Y. Simonin, D. van Riel, P. Van de Perre, B. Rockx, S. Salinas, Differential virulence between Asian and African lineages of Zika virus, PLoS neglected tropical diseases, 11 (2017) e0005821.

[2] E.B. Hayes, Zika virus outside Africa, Emerging infectious diseases, 15 (2009) 1347.

[3] F. Macnamara, Zika virus: a report on three cases of human infection during an epidemic of jaundice in Nigeria, Transactions of the royal society of tropical medicine and hygiene, 48 (1954) 139–145.

[4] G.W. Dick, S.F. Kitchen, A.J. Haddow, Zika virus (I). Isolations and serological specificity, Transactions of the royal society of tropical medicine and hygiene, 46 (1952) 509–520.

[5] E. Oehler, L. Watrin, P. Larre, I. Leparc-Goffart, S. Lastere, F. Valour, L. Baudouin, H. Mallet, D. Musso, F. Ghawche, Zika virus infection complicated by Guillain-Barre syndrome–case report, French Polynesia, December 2013, Eurosurveillance, 19 (2014).

[6] G. Calvet, R.S. Aguiar, A.S. Melo, S.A. Sampaio, I. De Filippis, A. Fabri, E.S. Araujo, P.C. de Sequeira, M.C. de Mendonça, L. de Oliveira, Detection and sequencing of Zika virus from amniotic fluid of fetuses with microcephaly in Brazil: a case study, The Lancet infectious diseases, 16 (2016) 653–660.

[7] M. Sarno, G.A. Sacramento, R. Khouri, M.S. do Rosário, F. Costa, G. Archanjo, L.A. Santos, N. Nery Jr, N. Vasilakis, A.I. Ko, Zika virus infection and stillbirths: a case of hydrops fetalis, hydranencephaly and fetal demise, PLoS neglected tropical diseases, 10 (2016) e0004517.

[8] F. Marinho, V.E.M.d. Araújo, D.L. Porto, H.L. Ferreira, M.R.S. Coelho, R.C.R. Lecca, H.d. Oliveira, I.P.d.A. Poncioni, M.H.N. Maranhão, Y.M.M.B. Mendes, Microcefalia en Brasil: prevalencia y caracterización de casos a partir del Sistema de Informaciones sobre Nacidos Vivos (Sinasc), 2000-2015, Epidemiologia e Serviços de Saúde, 25 (2016) 701–712.

[9] P.P. Garcez, E.C. Loiola, R. Madeiro da Costa, L.M. Higa, P. Trindade, R. Delvecchio, J.M. Nascimento, R. Brindeiro, A. Tanuri, S.K. Rehen, Zika virus impairs growth in human neurospheres and brain organoids, Science, 352 (2016) 816–818.

[10] F.R. Cugola, I.R. Fernandes, F.B. Russo, B.C. Freitas, J.L. Dias, K.P. Guimarães, C. Benazzato, N. Almeida, G.C. Pignatari, S. Romero, The Brazilian Zika virus strain causes birth defects in experimental models, Nature, 534 (2016) 267–271.

[11] P. Brasil, J.P. Pereira Jr, M.E. Moreira, R.M. Ribeiro Nogueira, L. Damasceno, M. Wakimoto, R.S. Rabello, S.G. Valderramos, U.-A. Halai, T.S. Salles, Zika virus infection in pregnant women in Rio de Janeiro, New England Journal of Medicine, 375 (2016) 2321–2334.

[12] W.H. Organization, Zika situation report: neurological syndrome and congenital anomalies, (2016).

[13] M.C. Castro, Q.C. Han, L.R. Carvalho, C.G. Victora, G.V. França, Implications of Zika virus and congenital Zika syndrome for the number of live births in Brazil, Proceedings of the National Academy of Sciences, 115 (2018) 6177–6182.

[14] F.A. Venancio, M.E. Quilião, D. de Almeida Moura, M.V. de Azevedo, S. de Almeida Metzker, L.K. Mareto, M.J. de Medeiros, C.D.B. Santos-Pinto, E.F. de Oliveira, Congenital anomalies during the 2015–2018 Zika virus epidemic: a population-based cross-sectional study, BMC Public Health, 22 (2022) 2069.

[15] M.P.A. Souza, M.S. da Natividade, G.L. Werneck, D.N. Dos Santos, Congenital Zika syndrome and living conditions in the largest city of northeastern Brazil, BMC Public Health, 22 (2022) 1231.

[16] B.L. de Amorin Vilharba, M. Yamamura, M.V. de Azevedo, W.d.S. Fernandes, C.D.B. Santos-Pinto, E.F. de Oliveira, Disease burden of congenital Zika virus syndrome in Brazil and its association with socioeconomic data, Scientific Reports, 13 (2023) 11882.

